# Seroprevalence of anti-SARS coronavirus 2 antibodies in Thai adults during the first three epidemic waves

**DOI:** 10.1101/2022.01.18.22269501

**Authors:** Hatairat Lerdsamran, Anek Mungaomklang, Sopon Iamsirithaworn, Jarunee Prasertsopon, Witthawat Wiriyarat, Suthee Saritsiri, Ratikorn Anusorntanawat, Nirada Siriyakorn, Poj Intalapaporn, Somrak Sirikhetkon, Kantima Sangsiriwut, Worawat Dangsakul, Suteema Sawadpongpan, Nattakan Thinpan, Pilailuk Okada, Ranida Techasuwanna, Noparat Mongkalangoon, Kriengkrai Prasert, Pilaipan Puthavathana

## Abstract

This study sought to determine the anti-SARS-CoV-2 antibody status of 4111 Thai people from May 2020 to April 2021, a period which spanned the first two and part of the third epidemic wave of the COVID-19 in Thailand. Participants comprised 142 COVID-19 patients, 2113 individuals at risk due to their occupations [health personnel, airport officers, public transport drivers, and workers in entertainment venues (pubs, bars and massage parlors)], 1856 individuals at risk due to sharing workplaces or living communities with COVID-19 patients, and 553 Thai citizens returning after extended periods in countries with a high disease prevalence. All sera were tested in a microneutralization assay and a chemiluminescence immunoassay (CLIA) for IgG against the N protein. Furthermore, we performed an immunofluorescence assay to resolve discordant results between the two assays. Antibody responses developed in 88% (15 of 17) of COVID-19 patients at 8 days and in 94-100% between 15 and 60 days after disease onset. Neutralizing antibodies persisted for at least 8 months, longer than the IgG did, against the N protein. None of the health providers, airport officers, and public transport drivers were seropositive, while the antibodies were present in 0.44% of entertainment workers. This study showed the seropositivity of 1.9, 1.5, and 7.5% during the 3 epidemic waves, respectively, in Bangkok residents who were at risk due to sharing workplaces or communities with COVID-19 patients. Also, antibody prevalence was 1.3% in Chiang Mai people during the first epidemic wave, and varied between 6.5 and 47.0% in Thais returning from high-risk countries. This serosurveillance study found a low infection rate of SARS-CoV-2 in Thailand before the emergence of the Delta variant in late May 2021. The findings support the Ministry of Public Health’s data, which are based on numbers of patients and contact tracing.

## Introduction

On 13 January 2020, Thailand was the first country to report a confirmed coronavirus disease-19 (COVID-19) case outside of China. The first indigenous case in Thailand occurred on 30 January 2020 in a local taxi driver who had no history of traveling abroad; investigation suggested that he was exposed to the severe acute respiratory syndrome coronavirus-2 (SARS-CoV-2) by a group of Chinese tourist passengers [1]. As of November 2021, Thailand had experienced 4 epidemic waves associated with different SARS-CoV-2 variants which emerged during the pandemic (Fig 1).

**Fig 1.**
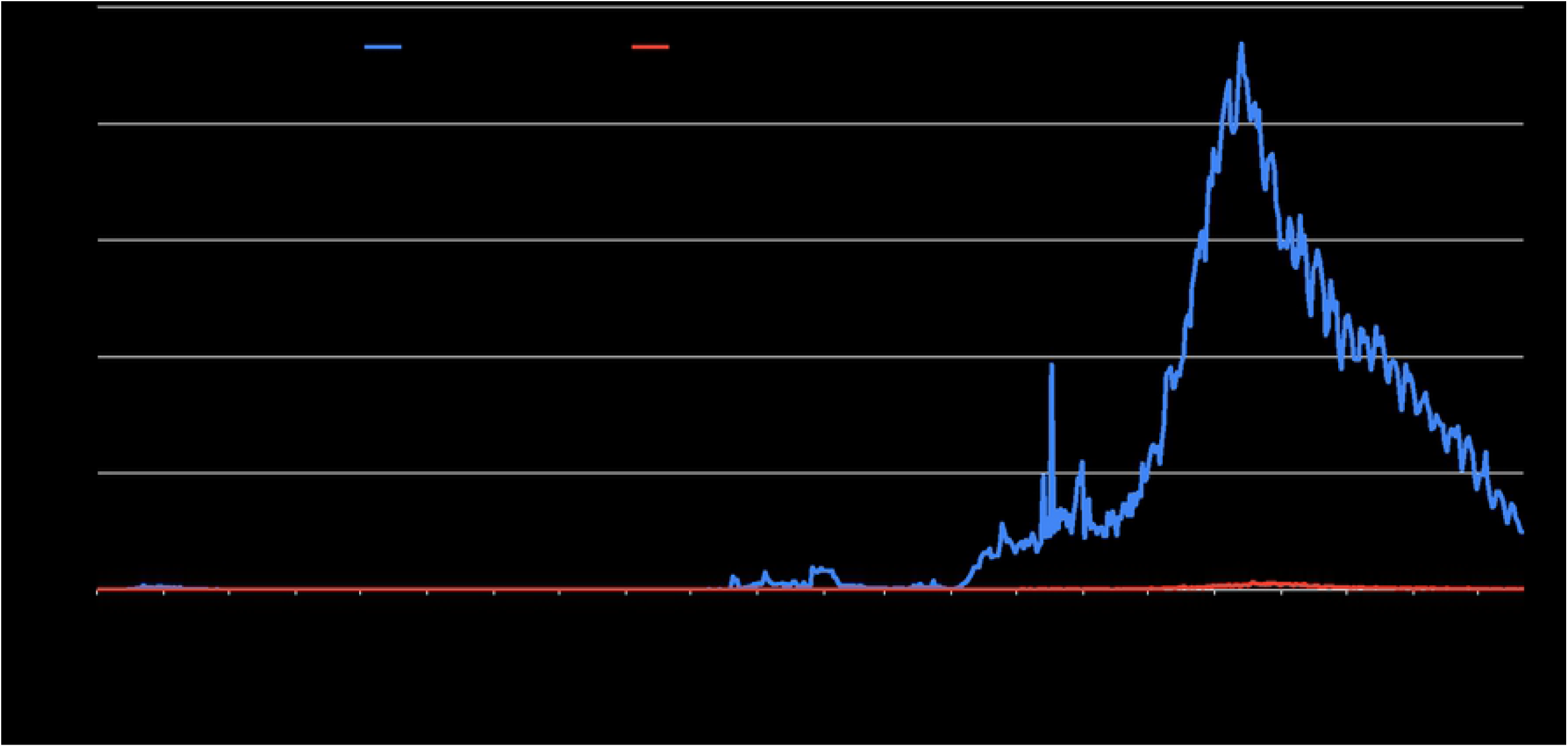
Situation of SARS-CoV-2 infection in Thailand during March 1, 2020 through December 21, 2021.

According to the Department of Disease Control (DDC), Ministry of Public Health (MOPH), the first epidemic wave began in January 2020, peaked in March-April, gradually declined, and ended on 14 December 2020. Large clusters occurring during this wave had linkages with attending boxing stadiums or entertainment venues in Bangkok. The strains causing this wave belonged to clades L, S, O, V, and G likely reflecting multiple introductions of virus into the country through tourism. The second wave was shorter, lasting from 15 December 2020 to 31 March 2021. This epidemic wave began among Myanmar migrant workers in a seafood market in Mahachai District, Samut Sakorn Province, 37 km south of Bangkok. The causative virus belonged to the GH clade (G614 mutation) which was circulating in Myanmar during the second epidemic wave since August 2020 [2]. The third epidemic wave in Thailand began on 1 April 2021, again in nightlife entertainment venues, and might be linked to the introduction of the Alpha variant (B.1.1.7) by the Thai people returning from Cambodia in the middle of March 2021 [3]. The Alpha variant had spread throughout Cambodia, particularly in Phnom Penh, since February 2021 [4]. The Alpha variant spread to many parts of Thailand in the second week of April due to movement of large numbers of people during the annual Songkran or water festival (the traditional Thai new year). Subsequently, the Delta variant, first detected among workers in a big construction camp at Laksi, Bangkok, on 7 May 2021 [5], spread countrywide seeding the fourth epidemic wave and caused loss of control of the epidemic.

People infected with SARS-CoV-2 may develop antibodies targeting multiple viral proteins, regardless of whether they have symptoms or not. Therefore, serosurveillance was an important tool for estimating the magnitude and monitoring the epidemic, especially since asymptomatic cases were common. A meta-analysis showed that the as many as 35% of cases were asymptomatic [6]. Serological data can also show the duration of antibody responses to COVID-19 infection, indicating partial protection from re-infection. Studies in Europe and the US have shown that neutralizing antibodies persist in a majority of COVID-19 cases for up to 13 months after infection [7], and patients with severe disease exhibited a higher level of neutralizing antibodies than those with milder disease symptoms [8]. The present study conducted a cross-sectional serosurveillance of anti-SARS-CoV-2 in Thai people in various risk groups in Bangkok and Chiang Mai Province from May 2020 to May 2021 (the duration spanned the first, second, and part of the third epidemic waves). The serological techniques used in this study included a microneutralization (microNT) assay, a chemiluminescence immunoassay (CLIA), and an indirect immunofluorescence assay (IFA).

## Material and methods

### Ethical statement

Usage of human sera from participants of age older than 18 received approval from the Mahidol University Central Institutional Review Board (MU-CIRB) under protocol number MU-COVID2020.001/2503.

### Study sites

Bangkok, the capital of Thailand, and Chiang Mai Province, 696 km north of Bangkok, were chosen as study sites due to their popularity as travel destinations, high population densities, and the high numbers of SARS-CoV-2-infected cases.

### Study population

The study involved 4111 serum samples from 4 groups of participants, as follows. 1) Anonymized archival serum samples from COVID-19 patients with no information of disease severity (Sera were the leftover samples from clinical laboratory investigations). 2) People at risk due to their occupations (health personnel, airport drivers, public transport drivers, and workers in entertainment venues (e.g., pubs, bars, and massage parlors). 3) People at risk due to sharing workplaces or communities with COVID-19 patients. In the enrollment process for participants in groups 2 and 3, epidemiologists explained the purpose of the study to obtain written consent for interviewing about their demographics, occupation, workplace, residence, and general health condition, including a donation of 5-8 ml of blood, with specimens labeled using ID codes. 4) Thai citizens in state quarantines, when arrived Thailand after extended duties in countries with known SARS-CoV-2 outbreaks. Blood specimens were collected for anti-SARS-CoV-2 antibody testing [along with real-time reverse transcription-polymerase chain reaction (RT-PCR)] to support active case surveillance activities conducted by the Institute for Urban Disease Control and Prevention (IUDC), DDC, MoPH. For this group, an ethical review was waived under the authorization of the DDC, MOPH, as part of the emergency public health response to the pandemic. Nevertheless, participants received explanations and gave verbal consent.

### Cell and virus cultivation

We cultivated Vero cells (African green monkey kidney cells - ATCC, CCL-81) in Eagle’s minimum essential medium (EMEM) (Gibco, NY) supplemented with 10% fetal bovine serum (FBS) (Gibco, NY), penicillin, gentamycin, and amphotericin B. SARS-CoV-2 was isolated and propagated in Vero cells maintained in EMEM supplemented with 2% FBS in a Biosafety Laboratory Level-3 facility. Using the Reed-Muench method, we calculated the virus concentration for 50% tissue culture infective dose (TCID50) for further use in the microNT assay.

### Test viruses

This study used 3 SARS-CoV-2 isolates as test viruses in the microNT assay. The purpose of doing so was to match the circulating virus at the time of blood specimen collection. The SARS-CoV-2 isolate designated hCoV-19/Thailand/MUMT-4/2020, clade O (GISAID accession number EPI_ISL_493139) was used as the test virus for sera collected during the first epidemic wave, while hCoV-19/Thailand/MUMT-13/2021, clade GH (GISAID accession number EPI_ISL_6267810), and hCoV-19/Thailand/MUMT-36/2021, clade GRY (B.1.1.7) (GISAID accession number EPI_ISL_6267895) were used as the test viruses for the second and third epidemic waves, respectively.

### Experimental design

This study employed the microNT assay and CLIA to detect anti-SARS antibodies in each test serum. When results of the two assays were concordant, the test serum was considered positive or negative for anti-SARS antibodies. In case of discordant results, the test serum was further investigated by IFA.

### Microneutralization assay

We used the cytopathic effect (CPE)-based MicroNT assay to determine levels of neutralizing (NT) antibodies against SARS-CoV-2 in the test sera. The assay employed Vero cell monolayers in 96-well microculture plates and SARS-CoV-2 at a final concentration of 100 TCID50/100 µl as the test virus. The method followed those described in our previous studies [9, 10]. Briefly, the test serum was heat-inactivated and serially two-fold diluted from 1:10 to 1:1280. A 60 µl volume of each serum dilution was mixed with 60 µl of the test virus at a concentration of 200 TCID50/100 µl. After an hour of incubation at 37 °C, a 100 µl volume of the virus-serum mixture was inoculated in duplicate into wells containing the Vero cell monolayer. The reaction plates were incubated at 37 °C for 3 days before reading the results. We defined the NT antibody titer as the highest reciprocal serum dilution that inhibited ≥50% CPE in the wells inoculated with the serum-virus mixture compared to wells with the uninfected cell control. A titer of 10 or greater was considered positive for NT antibody.

### Chemiluminescence immunoassay

CLIA using Architect auto-analyzer (Abbott Laboratories, USA) is the two-step, fully automated immunoassay that qualitatively detected binding between the SARS-CoV-2 nucleoprotein (N) antigen coated on paramagnetic microparticles and human IgG in the test sera. The assay required a minimal volume of 150 µl of test serum to fill the reaction cup. Acridinium-conjugated anti-human IgG bound the human IgG and then emitted chemiluminescence signals quantitated as a relative light units (RLUs). It took about 1 hour to complete a test run. The level of SARS-CoV-2 IgG was directly correlated to the amount of RLUs. An index value was established based on the ratio between the RLU of the kit positive sample (S) and the calibrator (C). A test serum with an S/C ratio ≥1.4 was considered positive for the SARS-CoV-2 IgG.

### Indirect immunofluorescence assay

The IFA staining method was previously described [10]. The assay employed SARS-CoV-2-infected Vero cells deposited on microscopic slides as the test antigens. To standardize the test antigen given the lot-to-lot variation, 50-75% of the infected cells must express N and spike (S1) proteins when stained with specific monoclonal antibodies (Sino Biological, Beijing, China). Human serum at a dilution of 1:10 in phosphate-buffered saline was incubated with the infected cell for 60 minutes at 37 °C and followed by addition of polyclonal rabbit anti-human IgA, IgG, IgM, Kappa, and Lambda conjugated with fluorescein isothiocyanate (Dako, Glostrup, Denmark) in Evan’s blue solution for 60 minutes. The stained viral antigens in the cytoplasm of the infected cells appeared apple green when examined under a fluorescence microscope.

### Statistical analysis

R square (R^2^), mean, and standard deviation (SD) were determined, and figures were drawn using GraphPad Prism version 8.4.3 for Windows (GraphPad Software, La Jolla, California, USA). The McNemar test was carried out and 95% confidence interval (95% CI) calculated by SPSS Statistic software version 18.0.

## Results

### Prevalence of anti-SARS-CoV-2 antibodies in COVID-19 patients by time

Using microNT assay and Architect IgG, and IFA for confirmation, anti-SARS-CoV-2 antibodies were detected in 124 (87.3%) of 142 COVID-19 patients from the first epidemic wave. Eighty-eight percent (15 of 17) of the patients had antibody responses at 8 days after onset of symptoms. Positivity rose further to 94.1% (16 of 17) at 15-21 days, 100% (19 of 19) at 22-30 days, and 97% (32 of 33) at 31-60 days after onset of symptoms (Fig 2). NT antibodies persisted for at least 8 months as found in all 7 participants with history of COVID-19, while IgG specific to N protein was found in only 3 of these 7 cases. Nevertheless, the numbers of antibody-positive cases detected by microNT assay and CLIA for IgG were not significantly different (McNemar test; *p* = 0.65). On the other hand, the NT antibody titers were not well correlate with the IgG levels (R^2^ = 0.6042) (Fig 3).

**Fig 2.**
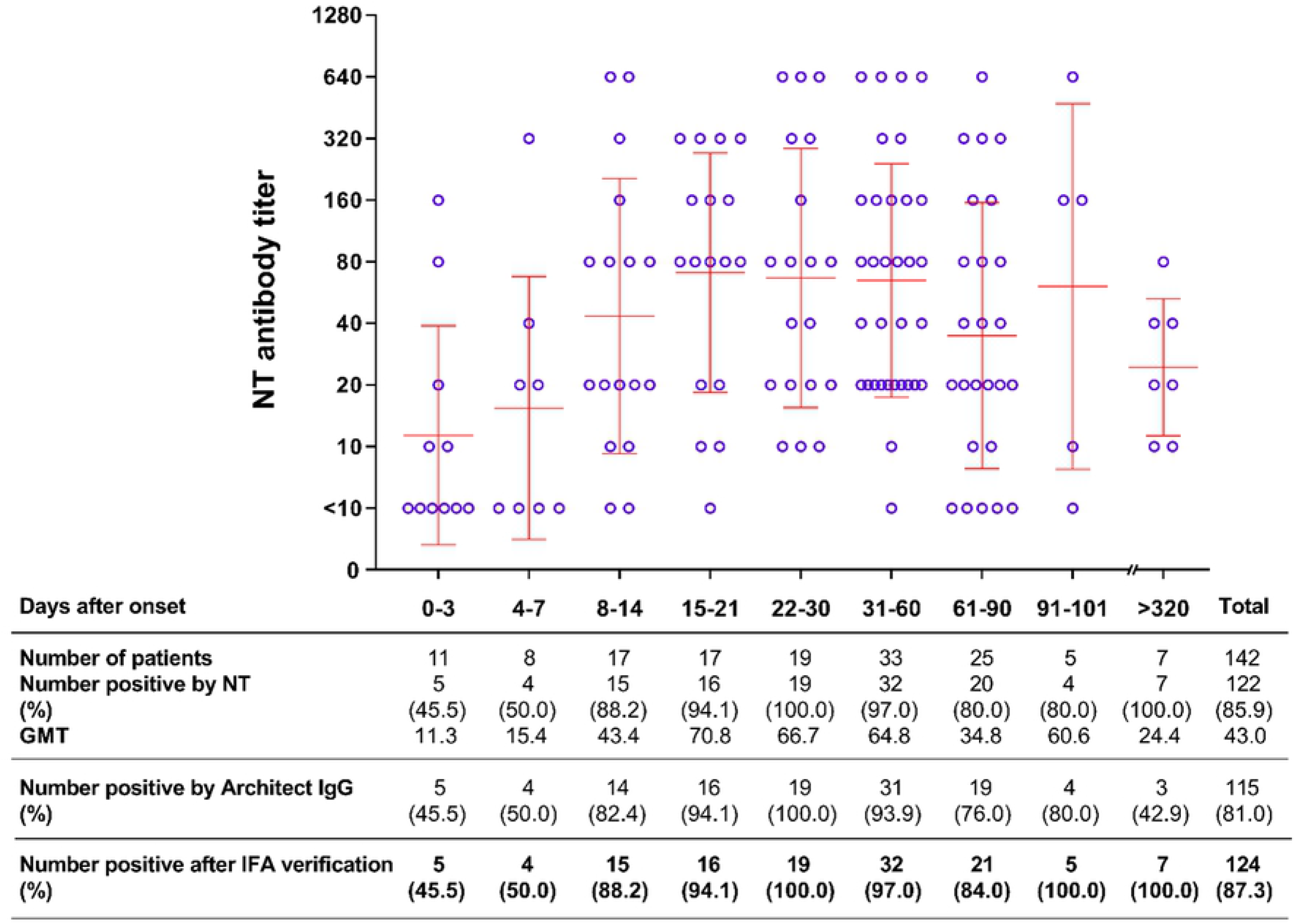
Antibody response in COVID-19 patients at different time points after onset of disease symptoms.

**Fig 3.**
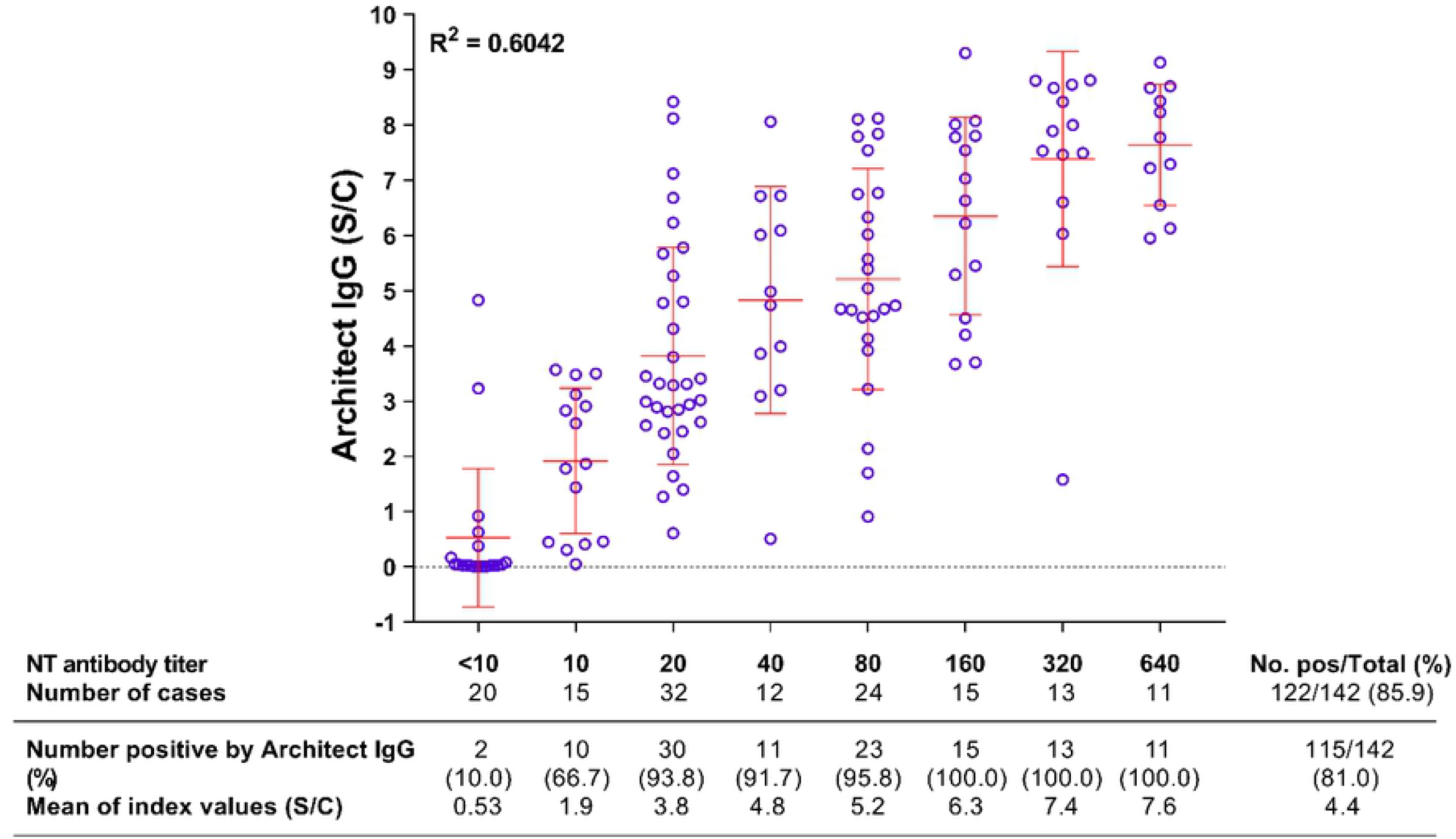
Correlation between NT antibody titers and Architect IgG indices in COVID-19 patients.

### Prevalence of anti-SARS-CoV-2 antibodies in participants with selected occupations

The early COVID-19 outbreak in Thailand showed that some occupations had higher risk of infection than did others. We conducted serosurveillance for anti-SARS-CoV-2 antibodies in 2113 participants with at-risk occupations (health personnel, airport officers, public transport drivers and workers in entertainment venues) as shown in Table 1. The serum samples from health personnel were collected during the three epidemic waves, while the others were collected from the first or second epidemic wave. The result showed that none of the 2113 participants had anti-SARS-CoV-2 antibodies, except for 3 (0.44%) of 682 workers from the entertainment venues.

**Table 1.** Seroprevalence of anti-SARS-CoV-2 antibodies in participants with at-risk occupations.

| Group | Epidemic wave |  |  | Subtotal |
| --- | --- | --- | --- | --- |
|  | Jan – December 14, 2020 | December 15, 2020 – March 31, 2021 | April 1 – May 31, 2021 |  |
|  | No. pos/<br>No. tested | No. pos/<br>No. tested | No. pos/<br>No. tested |  |
| Health personnel | 0/187 | 0/13 | 0/272 | 0/472 |
| Airport officers | 0/493 | - | - | 0/493 |
| Public transport drivers | 0/455 | 0/11 | - | 0/466 |
| Entertainment workers | 3/682 | - | - | 3/682 (0.44%)<br>95% CI = 0.08 – 1.29% |
| **Total = 3/2113 (0.14%), 95% CI = 0.03 – 0.42%** |  |  |  |  |

### Prevalence of anti-SARS-CoV-2 antibodies in people at risk

We conducted serosurveillance in 1856 Thai people at risk by sharing the same workplaces or living in the same community with COVID-19 cases. The investigation showed that 1.9% (11 of 574), 1.5% (6 of 388), and 7.5% (11 of 147) of people in Bangkok were seropositive for anti-SARS-CoV-2 antibody during the three epidemic waves, respectively. In Chiang Mai, 1.3% (10 of 747) of participants were seropositive during the first epidemic wave; 7 of them provided histories of having COVID-19 during the prior 8 months (Table 2).

**Table 2.** Seroprevalence of anti-SARS-CoV-2 antibodies in participants who shared workplaces or lived in communities with reported COVID-19 cases.

| Province | Epidemic wave |  |  |
| --- | --- | --- | --- |
|  | Jan – December 14, 2020 | December 15, 2020 – March 31, 2021 | April 1 – May 31, 2021 |
|  | No. pos/No. tested | No. pos/No. tested | No. pos/No. tested |
| Bangkok | 11/574 (1.9%)<br>95% CI = 0.95 – 3.43% | 6/388 (1.5%)<br>95% CI = 0.56 – 3.37% | 11/147 (7.5%)<br>95% CI = 3.72 – 13.40% |
| Chiang Mai | 10/747 (1.3%)<br>95% CI = 0.64 – 2.46% | - | - |
| **Subtotal** | 21/1321(1.6%)<br>95% CI = 0.98 – 2.43% | 6/388 (1.5)<br>95% CI = 0.56 – 3.37% | 11*/147 (7.5%)<br>95% CI = 3.72 – 13.40% |
| **Total = 38/1856 (2.04%), 95% CI = 1.44 – 2.81%** |  |  |  |

\* 7 participants provided history of COVID-19 at 8 months previously

### Serological profiles of anti-SARS-CoV-2 antibodies in participants at risk

We display in Tables 1 and 2 the serological profiles of anti-SARS-CoV-2 antibodies in 3969 participants, grouped as 2113 people at risk by occupation (Table 1), and 1856 people at risk by sharing workplaces or living community with COVID-19 patients. Using microNT assay and Architect IgG, followed by IFA validation, 41 of 3969 (1.0%) were seropositive for anti-SARS-CoV-2 antibodies. The number of seropositive cases detected by Architect IgG was slightly lower than that detected by microNT, but not significantly different (McNemar test; *p* = 0.54). Furthermore, the NT antibody titers were not well correlated with the IgG levels obtained by CLIA (R^2^ = 0.5908) (Fig 4).

**Fig 4.**
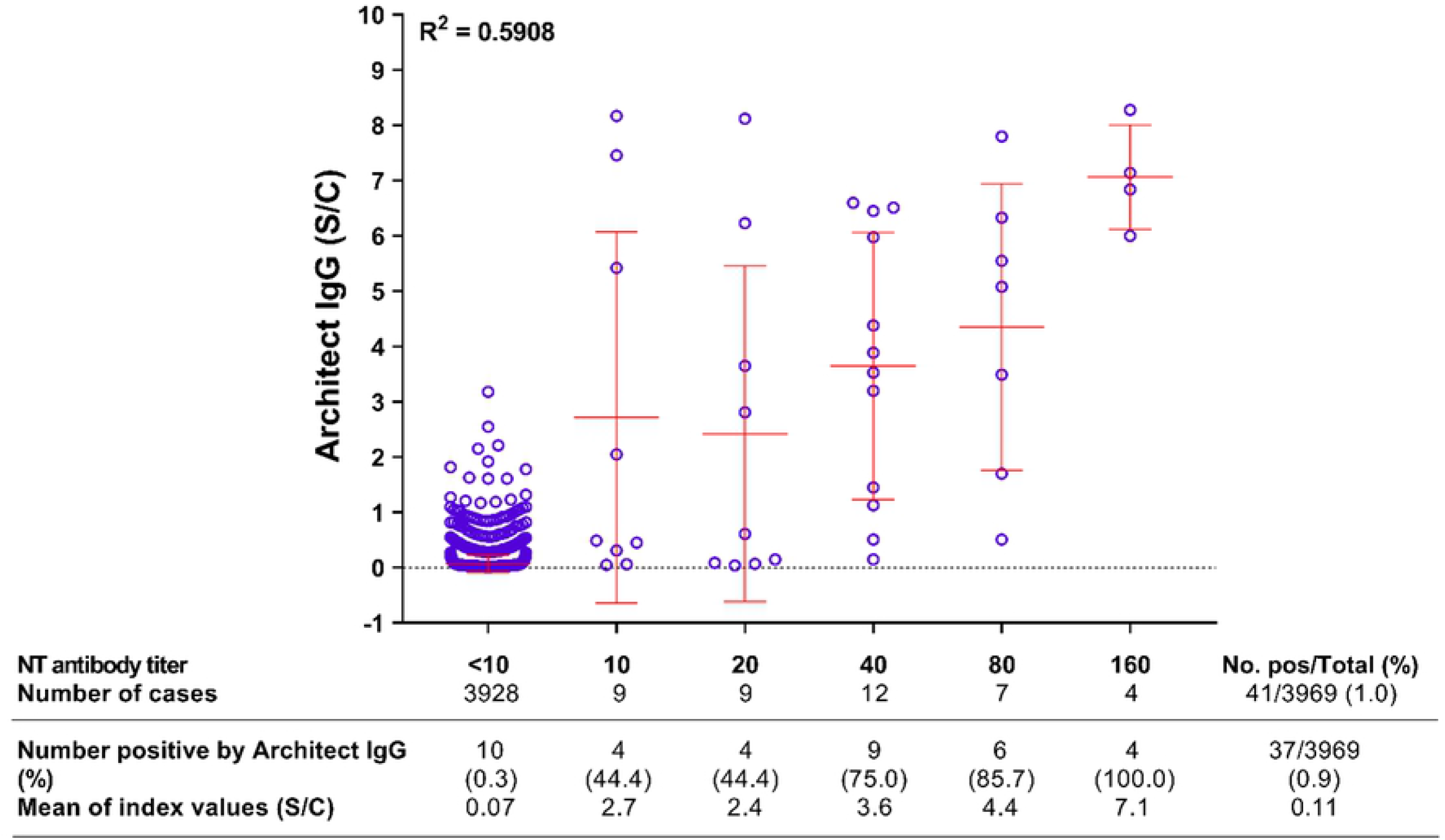
Correlation between NT antibody titers and Architect IgG indices in Thai people.

### Prevalence of anti-SARS-CoV-2 antibody in travelers returning from high-risk areas

This study conducted serosurveillance in 553 Thai citizens returning after extended duties in countries with high-prevalence of SARS-CoV-2 infection between May and October 2020. Our results showed seroprevalences of 6.5 – 47.0% depending on the country (Table 3).

**Table 3.** Seroprevalence of anti-SARS-CoV-2 antibodies in Thais returning from extended periods of work in high-risk countries.

| Country | Date of blood collection | No. pos/No. tested (%) |
| --- | --- | --- |
| Qatar | May – Jun 2020 | 14/215 (6.5%)<br>95% CI = 3.55 – 10.93% |
| Kuwait | May – Jun 2020 | 101/215 (47.0%)<br>95% CI = 38.27 – 57.08% |
| Sudan | Oct 2020 | 36/77 (46.8%)<br>95% CI = 32.75 – 64.72% |
| Others | Jun – Oct 2020 | 4/46 (8.7%)<br>95% CI = 2.28 – 22.36% |
| **Total = 155/553 (28.0%), 95% CI = 23.79 – 32.80%** |  |  |

## Discussion

We employed 3 serological techniques to detect anti-SARS-CoV-2 antibodies in this cross-sectional surveillance study. The NT antibodies were directed against the neutralizing epitopes present in the receptor-binding domain (RBD), N terminal domain (NTD), and S2 domain in the S protein [11–15]; Architect IgG was directed against the N antigen which is a more conserved protein [16, 17]. NT antibodies are markers of protective immunity, but IgG against N protein is not. MicroNT assay and Architect IgG were used to investigate every serum sample, and the result was concluded to be negative when both methods yielded concordant results. In the case of discordant results, IFA (which employed the test antigens from both the S and N proteins) was used to resolve discordance. Overall, the numbers of seropositive participants determined by microNT assay and Architect IgG were not significantly different. Nevertheless, NT antibody titers were not well correlated with the IgG levels.

Our study in COVID-19 patients showed that 15 of 17 (88.2%) patients mounted a detectable antibody response 8-14 days after onset of symptoms. However, the prevalence increased to as high as 94-100% in the subsequent 45 days. In addition, we found that anti-SARS-CoV-2 antibodies persisted for at least 8 months in all 7 individuals who had a history of COVID-19, while Architect IgG to N protein did not last that long. This is similar to the findings of others who reported 8-month antibody persistence in mild SARS-CoV-2 infections [18] and up to one year in one study [19].

We investigated 3969 blood samples from multiple groups of participants between May 14, 2020 and May 21, 2021, the duration spanning two epidemic waves and part of the third wave. Of 2113 participants with the at-risk occupations (472 health providers, 493 airport officers, 466 public transport drivers, and 682 workers in entertainment venues), only 0.14% (all 3 from the last group) had detectable anti-SARS-CoV-2 antibodies. Furthermore, of 1856 participants who shared workplaces or communities, only 38 (2.04%) were seropositive. Over time, the number of participants in Bangkok who had anti-SARS-CoV-2 antibodies increased from 1.9 to 1.5 and 7.5% during the 3 epidemic waves, respectively. Our seroprevalence data strongly supported the prevalence of infection reported by the MoPH, i.e., the cumulative numbers of 4237 cases with 60 deaths at the end of the first epidemic wave (from Jan to Dec 14, 2020), 28863 cases with 94 deaths at the end of the second epidemic wave (from Dec 15, 2020 to Mar 31, 2021), and 159792 cases with 1031 deaths (from Apr 1 to May 31, 2021).

Thailand received worldwide recognition for keeping the first epidemic wave well controlled. During the first epidemic wave, cumulative case count per population was in the 10^th^ percentile of countries globally [20]. As part of the control policy, the government provided treatment and hospitalization, quarantine, and laboratory diagnosis of SARS-CoV-2 infection at no cost. The prevalence of infection during the wave peaked in March 2020, followed by a sharp decline. None or less than 10 locally transmitted cases were detected daily from the middle of May through the end of the first epidemic wave. In total, there were 4237 cases with 60 deaths by the end of the first epidemic wave (December 14, 2020). The high seroprevalence (6.5 – 47%) in Thai citizens returning from abroad suggested the seriousness of the outbreaks in those regions. Nevertheless, high infection rates may be partly due to sharing workplaces among these participants.

Management of non-pharmaceutical interventions in Thailand was efficient. No people protested against wearing masks in public. They followed the suggestions on soft locked down, social distancing, working from home, and personnel hygiene. A vital success came from the assistance of approximately one million village health volunteers who are part of the public health system and worked nationwide since the time of H5N1 avian influenza. These volunteers assist with health education, active case finding, and communication between health authorities and communities. For example, each volunteer is assigned to take care of approximately 10 houses. Nevertheless, the occurrence of the second and third epidemic waves came very abruptly, from introduction of the newer variants, the GH clade and the Alpha variant, respectively. We cannot deny that these outbreaks due to illegal activities, including cross border movement of migrant workers and gamblers.

Before the third epidemic wave trended down, the outbreak situation of SARS-CoV-2 became worse due to introduction of the Delta variant (that is more transmissible and virulent) to seed a fourth epidemic wave. The infection rate peaked in August 2021, when more than 20000 cases were reported daily for weeks before slowly declining. However, non-pharmaceutical intervention appears inadequate to control the Delta variant, and access to vaccines has not been easy. The MoPH first launched vaccinations for health providers in late February 2021.

At present, seroprevalence studies in the Thai population will encounter difficulties in differentiating between natural infection and vaccination. Nevertheless, our seroprevalence data validate the reported lack of circulation of SARS-CoV-2 in Thailand during the first year of the pandemic.

## Data Availability

All relevant data are within the manuscript.

## Acknowledgements

We thank Abbott Laboratories for providing Chemiluminescence test kits. We also thank Chinda Kanoksinsombat, Tipsuda Chanmanee, Rumporn Kularb, and Charlearnsak Thapyotha for laboratory assistance.

